# The Prognostic Respiratory Intensity Scoring Metric (PRISM)

**DOI:** 10.1101/2025.06.28.25330368

**Authors:** Madelyn G. Nance, Carrington S. Davis, Zoe G. Kitchings, Chad Aldridge, Santina Zanelli, Jennifer Burnsed, Meghan H. Puglia

## Abstract

Invasive respiratory interventions save infant lives yet, have negative consequences for their general health and neurodevelopment. Tools that can both accurately summarize the intensity of ventilation needed during an infant’s stay in the NICU and be used to make predictions about their future health and neurodevelopmental outcomes are currently lacking. Here we present the Prognostic Respiratory Intensity Scoring Metric (PRISM) as an accurate, summative tool of respiratory support needs in hospitalized neonates well suited for predicting an infant’s future neurodevelopmental outcomes. 218 infants were included in this study. We trained a classification and regression tree model using PRISM scores to classify respiratory diagnoses and compared performance to the Neonatal Sequential Organ Failure Assessment Respiratory subscore (nSOFA-r). PRISM outperforms the nSOFA-r in classifying infants with respiratory distress syndrome (PRISM AUC = 0.71, nSOFA-r AUC = 0.64) and chronic lung disease (PRISM AUC = 0.86, nSOFA-r AUC = 0.76) according to Delong’s Test (p=0.002, p=0.001). PRISM had significant associations both with an infant’s length of hospital stay (β = 0.03 (95% CI [0.02, 0.03], p < 0.001) and gestational age (β =−0.03 (95% CI [−0.03, −0.02], p <0.001). PRISM successfully identifies which infants will develop an intraventricular hemorrhage (83% accuracy) and is highly successful at determining which infants will develop retinopathy of prematurity (95% accuracy). PRISM is a tool designed for clinicians and researchers alike to summarize the invasive nature of neonatal respiratory support.

## 1. Introduction

When an infant is born premature, or before 37 weeks gestational age^1^ (GA), lung maturation is disrupted^2,3^. As a result, many premature infants are diagnosed with respiratory distress syndrome (RDS) at birth, often necessitating invasive mechanical ventilation^4^. If respiratory support and/or supplemental oxygen are still required at 36 weeks corrected GA a diagnosis of bronchopulmonary dysplasia (BPD)^5^ or chronic lung disease (CLD)^6^ is made. BPD and CLD are both associated with chronic respiratory dysfunction^7,8^ and adverse neurodevelopmental outcomes^9,10^. In 2023, over 10% of infants in the United States were born preterm^11^ and up to 85% of those infants required some form of respiratory intervention^12^ while in the Neonatal Intensive Care Unit (NICU). Given these high prevalence rates, understanding the cumulative effect of respiratory intervention and its long term implications for the developing child is a significant public health goal.

Modes of respiratory support in the NICU range from non-invasive techniques such as continuous positive airway pressure (CPAP) or non-invasive positive pressure ventilation (NIPPV) to invasive forms of mechanical ventilation. While mechanical ventilation saves lives it is also associated with adverse health outcomes including the development of BPD/CLD^13,14^, and is associated with the exacerbation of retinopathy of prematurity^15,16^ (ROP), and intraventricular hemorrhage^17,18^ (IVH). In addition, prolonged need for mechanical ventilation is associated with adverse neurodevelopmental outcomes^9,10,19,20^, meaning that avoiding intubation and focusing on non-invasive methods of respiratory support is an important management consideration when caring for preterm infants. At present, clinicians lack tools to accurately represent the degree of respiratory support that infants experience throughout their stay in the NICU.

In recent decades, our understanding of the long term consequences ventilators have on health and neurodevelopment has become increasingly clear^9,10,19,20^, and recent guidelines suggest the use of less invasive forms of ventilation^7,21,22^, although with mixed results^20^. Some clinical decision-making tools exist to help doctors assess an infant’s immediate respiratory needs^22,23^ or predict the incidence of severe respiratory outcomes such as BPD or death^24^. One such tool, the Neonatal Sequential Organ Failure Assessment^25^ (nSOFA), generates a measure of lung function in mechanically ventilated patients^26^ through the respiratory subscore (nSOFA-r). Scores range from 0 to 8 and are calculated from the ratio of peripheral oxygen saturation to the fraction of inspired oxygen. However, the nSOFA-r is a fairly new score and has not yet been thoroughly tested as a respiratory indicator. To our knowledge, no scoring tool has been developed to effectively summarize the entirety of an infant’s ventilatory experience in the NICU in order to eventually facilitate the prediction of their neurodevelopmental outcomes.

The goal of this study was to develop a summative tool, the Prognostic Respiratory Intensity Scoring Metric (PRISM), capable of accurately representing levels of respiratory intervention required by an infant during their hospital stay, with the ultimate aim of using it to predict neurodevelopmental outcomes. We demonstrate PRISM’s use as a holistic measure of an infant’s respiratory experience in the NICU by establishing its ability to discriminate between respiratory diagnoses and incidences of respiratory comorbidities (ROP and IVH). PRISM is designed for clinicians and researchers alike to summarize invasive respiratory support and provides an optimal metric for predicting an infant’s future neurodevelopmental outcomes.

## 2. Methods

### 2.1. Participant Population

This is a secondary data analysis from a longitudinal, neurodevelopmental parent study. In the parent study, infants are recruited from the UVA NICU or the greater Charlottesville area. Infants recruited in the NICU are referred to the study team by neonatologists who determine the child’s health is sufficiently stable for participation. Infants are excluded if their mother is Non-English-speaking, a prisoner, unable to return for longitudinal follow ups, or if the infant has an uncorrectable severe auditory or visual deficit. Given our need to extract detailed respiratory information from electronic health records for the present study, only infants who received care at the UVA hospital (n=218) are included in this analysis.

### 2.2. PRISM Score Calculation

Custom R^27^ scripts extract each type of ventilation, ventilator settings, and nSOFA-r score from each infant’s EHR on each day of their hospitalization. To assign each infant a PRISM score representing their entire course of respiratory support in the NICU, we begin by evaluating the resuscitation required in the delivery room. Delivery room resuscitation methods were categorized into 9 different delivery room resuscitation types (Table 1) based on the intensity of required support. The infant was assigned a delivery room score corresponding to the most invasive delivery room resuscitation type received at birth Next, we considered ongoing respiratory support throughout the hospital stay. Following an infant’s discharge from the hospital, we extracted the most frequently charted ventilation type for each day of the hospitalization, and then summed the number of consecutive days the infant received that form of ventilation to determine its duration. Ventilation types were categorized based on the invasiveness of the respiratory support and assigned a score (Table 2). For each episode, the ventilation type was multiplied by the episode’s duration to generate a ventilation score. For each infant, all ventilation scores were summed and then added to the delivery room score to generate a final PRISM score. For each infant, extracted nSOFA-r scores were similarly summed across their entire stay in the hospital to generate a comparable, comprehensive, and summative score. In exploratory analyses, we also computed PRISM over the first 24 hours (PRISM-1) and 5 days of an infant’s hospital stay (PRISM-5) to determine their ability to predict respiratory diagnoses early in life. The results of analyses using these alternate PRISM scores are presented in the Supplemental Materials. The intensity and invasiveness of each Delivery Room Resuscitation Method and Ventilation Method were categorized, ranked, and assigned a corresponding score in collaboration with a consulting neonatologist (JB) such that the highest scores are assigned to the most invasive methods.

**Table 1:** Delivery Room Score and Ventilation Score Assignments.

| Score Type | Assigned Score | Ventilation / Resuscitation Method |
| --- | --- | --- |
| Delivery Room Scores | 9 | **Resuscitative Medications** |
|  | 8 | **Chest Compression** |
|  | 7 | **Intubation** |
|  | 6 | **Positive Pressure Ventilation** |
|  | 5 | **Continuous Positive Airway Pressure** |
|  | 4 | **Supplemental Oxygen** |
|  | 3 | **Deep Suction** |
|  | 2 | **Suction** |
|  | 1 | **Stimulation** |
| Ventilation Scores | 7 | **High Frequency Ventilation**<br>(High Frequency Jet Ventilation, High Frequency Oscillatory Ventilation) |
|  | 6 | **Conventional Ventilation**<br>(Drager Ventilation, Conventional Ventilation, Pressure Controlled/ Pressure Supported Ventilation, Assist Control Ventilation with Volume Guarantee and Pressure Support, Synchronized Intermittent Mandatory Ventilation, Volume Targeted Ventilation, Mechanical Ventilator) |
|  | 5 | **Non-Invasive Ventilation**<br>(Non-Invasive Positive Pressure Ventilation, Non-Invasive Ventilation) |
|  | 4 | **Continuous Positive Airway Pressure (CPAP)**<br>(Variable CPAP, Bubble CPAP, Positive-End-Expiratory Pressure) |
|  | 3 | **High Flow Nasal Cannula** |
|  | 2 | **Low Flow Nasal Cannula** |
|  | 1 | **Room Air** |

**Table 2:** Demographic, Medical, Obstetric characteristics of our patient population.

| Degree of Prematurity |  |
| --- | --- |
| GA (weeks), Mean±SD | 32.4 ± 3.9 |
| Extremely Preterm (<28 weeks) | 38 (17%) |
| Very Preterm (28-32 weeks) | 39 (18%) |
| Moderate Preterm (32-34 weeks) | 71 (33%) |
| Late preterm (34-37 weeks) | 46 (21%) |
| Early Term (37-39 weeks) | 16 (7%) |
| Full Term (39-41 weeks) | 7 (3%) |
| Late Term (>41 weeks) | 1 (<1%) |
| Multiplicity |  |
| Singleton | 162 (74%) |
| Twin | 53 (24%) |
| Triplet | 3 (2%) |
| Sex |  |
| Female | 106 (49%) |
| Male | 112 (51%) |
| Race |  |
| White | 152 (70%) |
| Black | 37 (17%) |
| More than 1 Race | 11 (5%) |
| Other | 18 (8%) |
| Ethnicity |  |
| Hispanic | 18 (8%) |
| Not Hispanic | 199 (91%) |
| Unknown | 1 ( 1%) |
| Birth Method |  |
| Vaginal | 71 (33%) |
| Cesarean Section | 147 (67%) |
| Respiratory Diagnoses |  |
| No Diagnosis | 44 (20%) |
| Respiratory Distress Syndrome | 119 (55%) |
| Chronic Lung Disease | 55 (25%) |
| Neonatal Medical Complications |  |
| Retinopathy of Prematurity | 69 (32%) |
| Intraventricular Hemorrhage | 29 (13%) |
| Pneumothorax | 6 (3%) |
| Hemodynamically Significant Patent Ductus Arteriosus or Patent Foramen Ovale | 8 (4%) |
| Hypoxic Ischemic Encephalopathy | 1 (0.5%) |

Two researchers (ZGK, CSD) identified diagnoses related to an infant’s respiratory function through a manual search for RDS or CLD/BPD in the infant’s discharge summary and list of diagnoses. If neither RDS nor CLD were listed in an infant’s chart in either location they were recorded as having no respiratory diagnosis (NoDx). Results were compared to an independent researcher (MGN) who manually recorded diagnoses for 130 of the 218 included infants with 100% agreement between these two independent queries. For our purposes at UVA, CLD and BPD are used interchangeably and for the remainder of this study CLD captures both diagnoses. The two researchers also manually recorded other common neonatal medical conditions such as ROP and IVH.

### 2.3. Assessing PRISM Score Performance

We assessed PRISM’s ability to classify the most commonly occurring respiratory disorders in the NICU. Using the caret package^28^ in R^27^, we first set a consistent seed value to ensure that random data handling conducted throughout analysis would be reproducible. Next, we randomly split 80% of the data into a training set (n=175) and the remaining 20% of the data was used to test model performance (n=43). A classification and regression tree (CART) model^29^ was built using the recursive partitioning method (rpart) from the caret package^28^ in R^27^ to classify infants into respiratory diagnosis groups based on their PRISM score. The model was trained using 5 fold cross validation to avoid overfitting. The complexity parameter was tuned across 10 candidate values, and all training and tuning was performed on the training dataset.

Next, predicted class probabilities were generated for both the training and test datasets.

We determined the optimal classification thresholds for each diagnostic class by computing Youden’s J statistic (determines the optimal threshold with equal weight given to sensitivity and specificity^30^) from the Receiver Operating Characteristic (ROC) curve of each class (generated using a one vs. all approach). We then performed predictions on the test set by comparing the predicted probabilities against their ROC-optimized classification thresholds. If a class probability exceeded its assigned threshold, the class with the highest predicted probability was assigned. An identical CART model was developed using nSOFA-r scores to compare model performance. To assess PRISM’s ability to represent the potential consequences of extended respiratory support, we also trained CART models (identical to those used to classify respiratory diagnoses) to classify the diagnosis of ROP and IVH based on an infant’s PRISM score.

We assessed the performance of each CART model by calculating the accuracy (fraction of correct classifications over total classifications), precision (ratio of true positives to all identified positives), recall (sensitivity / correct identification of a true positive) and F1 score (measure of accuracy from precision and recall) for each possible class^35^ along with the micro and macro-average F1 score, precision, and recall (Table 3).

**Table 3:** Model Fit Statistics.

|  | Threshold | Precision | Recall | F1 Score | Accuracy |
| --- | --- | --- | --- | --- | --- |
|  | PRISM Score |  |  |  |  |
| No Diagnosis | 0.15 | 1.00 | 0.24 | 0.38 | 74% |
| Respiratory Distress Syndrome | 0.38 | 1.00 | 0.42 | 0.59 |  |
| Chronic Lung Disease | 0.47 | 0.91 | 1.00 | 0.95 |  |
| Micro-Average |  | 0.94 | 0.56 | 0.7 |  |
| Macro-Average |  | 0.97 | 0.56 | 0.64 |  |
|  | AUC |  |  |  |  |
|  | PRISM Score |  |  |  |  |
| No Diagnosis | 0.60 [0.56, 0.65] |  |  |  |  |
| Respiratory Distress Syndrome | 0.71 [0.65, 0.76] * |  |  |  |  |
| Chronic Lung Disease | 0.86 [0.80, 0.92] * |  |  |  |  |
| Micro-Average | 0.84 [0.80, 0.88] |  |  |  |  |
| Macro-Average | 0.72 [0.68, 0.76] |  |  |  |  |
|  | Threshold | Precision | Recall | F1 Score | Accuracy |
|  | nSOFA-r Score |  |  |  |  |
| No Diagnosis | 0.14 | 1.00 | 0.21 | 0.34 | 62% |
| Respiratory Distress Syndrome | 0.35 | 0.71 | 0.26 | 0.39 |  |
| Chronic Lung Disease | 0.51 | 0.83 | 0.94 | 0.88 |  |
| Micro-Average |  | 0.84 | 0.49 | 0.62 |  |
| Macro-Average |  | 0.85 | 0.47 | 0.54 |  |
|  | AUC |  |  |  |  |
|  | nSOFA-r Score |  |  |  |  |
| No Diagnosis | 0.59 [0.55, 0.62] |  |  |  |  |
| Respiratory Distress Syndrome | 0.64 [0.60, 0.69] |  |  |  |  |
| Chronic Lung Disease | 0.76 [0.69, 0.83] |  |  |  |  |
| Micro-Average | 0.79 [0.74, 0.84] |  |  |  |  |
| Macro-Average | 0.66 [0.62, 0.71] |  |  |  |  |
|  | Threshold | Precision | Recall | F1 Score | Accuracy |
| Intraventricular Hemorrhage |  |  |  |  |  |
| Yes | 0.16 | 0.97 | 0.84 | 0.90 | 83% |
| No | 0.84 | 0.40 | 0.80 | 0.53 |  |

**Table 3: Model Fit Statistics.**
| Retinopathy of Prematurity |  |  |  |  |  |
| --- | --- | --- | --- | --- | --- |
| Yes | 0.48 | 0.94 | 1.00 | 0.97 | 95% |
| No | 0.52 | 1.00 | 0.85 | 0.92 |  |

To visualize the diagnostic performance of the PRISM and nSOFA-r models and demonstrate the balance between sensitivity and specificity across various thresholds, we conducted an ROC analysis for the PRISM and nSOFA-r models model using the pROC^36^ package in R^27^. These models have multi-class outcomes and as such, we constructed one-vs-all binary outcomes for each class (RDS, CLD, NoDx) as well as the micro-average and macro-average ROC curves. For each ROC curve, we calculated the area under the curve (AUC) values with a 95% confidence interval (CI) using a 10,000 iteration bootstrap resampling technique. Micro and macro-average AUCs are useful when evaluating the performance of a multi-class identifier because they represent both a global AUC and an average of each binary AUC respectively^37^. For each binary class outcome we used DeLong’s test^38^ to determine whether the AUCs using PRISM were significantly different from AUCs using the nSOFA-r.

We hypothesized that if PRISM is a reliable measure of the intensity of an infant’s need for respiratory support and their overall respiratory health, it would be significantly associated with the length of their hospital stay and their GA, as both of these variables are associated with an infant’s need for ventilation^31–33^. To test this hypothesis, we ran two separate gamma regressions with a log link to assess the relationship between each infant’s PRISM score and their length of stay and GA, respectively. We chose gamma regression since PRISM values are all positive and have a right skew^34^. Statistical Analysis

## 3. Results

### 3.1. Participant characteristics

Data were extracted from the electronic health records (EHR) of 218 infants who received care in the University of Virginia NICU and participated in an ongoing longitudinal neurodevelopment study. Demographic, medical, and general obstetric information was collected from each infant (Table 2).

### 3.2. PRISM scores can effectively classify respiratory diagnoses and outperform the nSOFA-r score

A CART model was trained on PRISM scores to predict respiratory diagnoses. This model achieved 74% accuracy overall, 73% accuracy in identifying CLD, 100% accuracy in identifying RDS, and 0% accuracy identifying NoDx (Figure 1A). The highest F1 score was for CLD (Table 3). ROC curves were plotted to visualize model performance (Figure 1B) and an AUC was calculated for each curve (Table 3). CLD had the highest AUC, followed by RDS and NoDx. The AUC for CLD and the Micro-average were well above the threshold for clinical utility.

**Figure 1:**
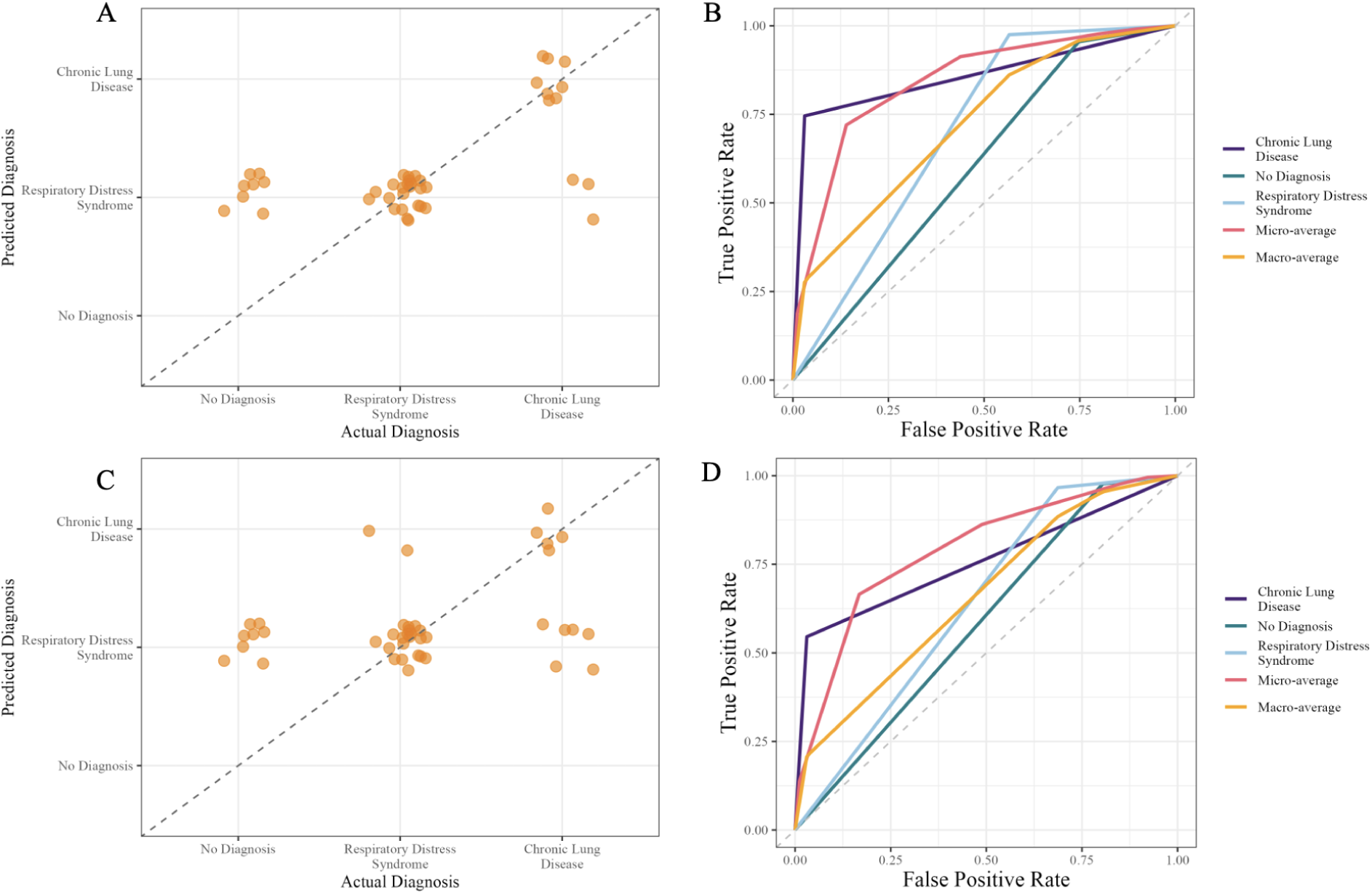
PRISM scores outperform the nSOFA-r score to discriminate between infants with the most common respiratory disorders in the NICU. **A.** A CART model was trained on PRISM scores to predict an infant’s respiratory diagnosis. A scatter plot of predicted and actual diagnosis visualizes the 74% accuracy of the model, with the highest accuracy for CLD. **B.** The performance of the CART model trained with PRISM to classify RDS, CLD, and NoDx as well as the micro and macro-average curves were visualized with an ROC analysis. The AUCs for all of these classes are included in Table 3. **C.** A CART was trained on nSOFA - r scores to predict an infant’s respiratory diagnosis. A scatter plot of predicted and actual diagnosis demonstrates the 62% accuracy for the model. **D.** The performance of the CART to classify RDS, CLD, and NoDx as well as the micro and macro-average curves were visualized with an ROC analysis. The AUCs for all of these classes are included in Table 3.

The same classifier was trained with nSOFA-r scores as a predictor. We found that nSOFA-r correctly predicted an infant’s respiratory diagnosis with 62% accuracy overall, 46% accuracy for CLD, 91% accuracy for RDS, and 0% accuracy for No Diagnosis (Figure 1C).

Again CLD had the highest F1 Score (Table 3). ROC curves were plotted (Figure 1D) and the AUC calculated for each curve (Table 3). CLD had the highest AUC followed by RDS and NoDx. The AUC for CLD and the Micro-average fall right at the threshold for clinical utility but are still lower than those of PRISM.

A Delong’s test of correlated ROCs determined that PRISM outperformed nSOFA-r in discriminating between respiratory diagnoses with significantly higher AUCs for CLD and RDS respectively (p < 0.001 and p = 0.002)

### 3.3. PRISM scores are highly accurate at discriminating between infants who develop or do not develop intraventricular hemorrhage and retinopathy of prematurity

Two CART models were trained to classify infants as having ROP and IVH respectively.

IVH was detected in the test set with 83% accuracy (Figure 2A) and ROP was detected with 95% accuracy (Figure 2B). The Precision, Recall, and F1 Score of each model can be seen in Table 3.

**Figure 2.**
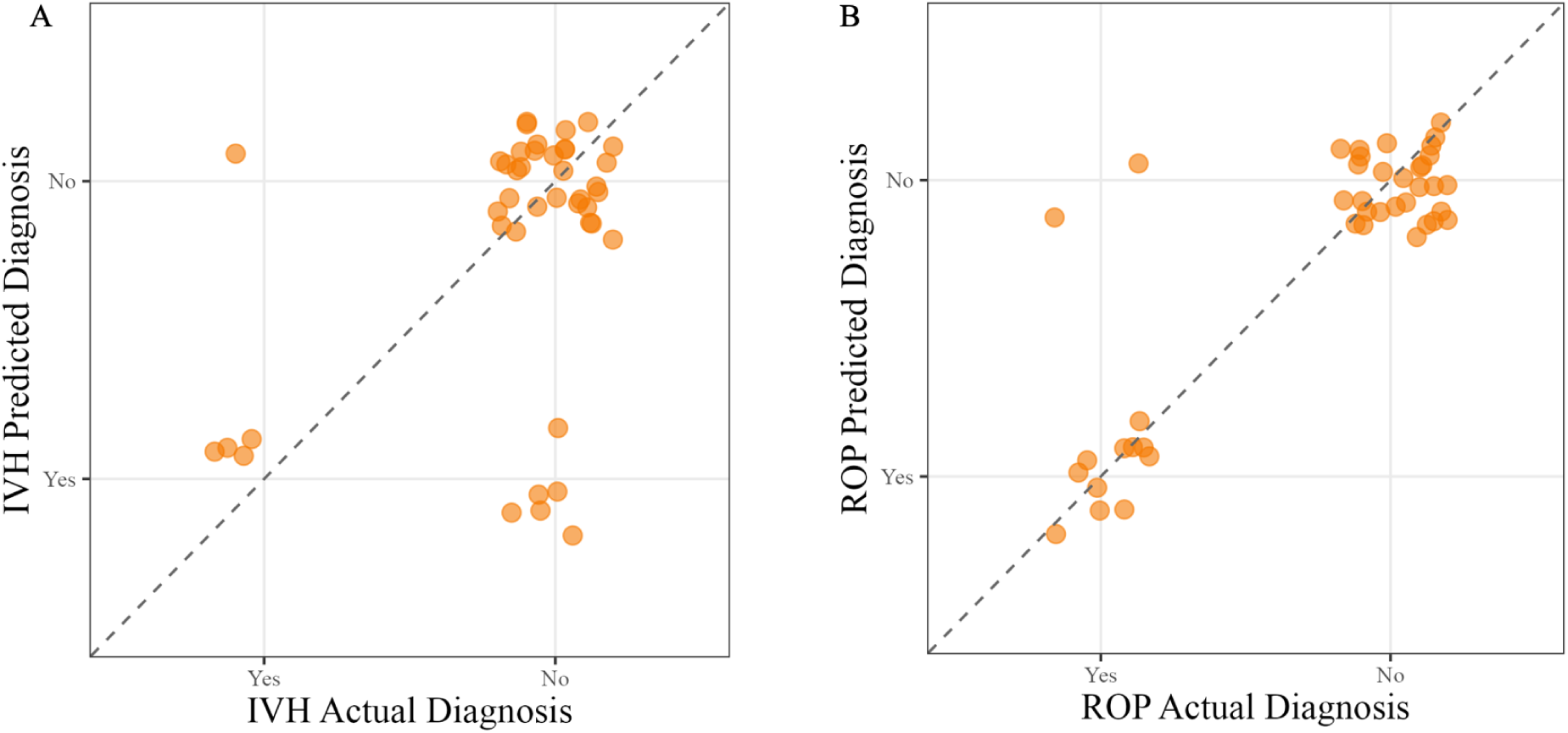
PRISM Scores can accurately distinguish between infants who do and do not develop both IVH and ROP. **A.** A CART model was trained on PRISM scores to predict an infant’s IVH diagnosis. A scatter plot of predicted and actual diagnosis visualizes the 83% accuracy of the model. **B.** A CART model was trained on PRISM scores to predict an infant’s ROP diagnosis. A scatter plot of predicted and actual diagnosis visualizes the 95% accuracy of the model.

### 3.4. PRISM scores are significantly associated with gestational age and hospital length of stay

We found a significant positive relationship between PRISM and length of hospital stay (β = 0.03 (95% CI [0.02, 0.03), p < 0.001) where every additional day in the hospital is associated with a 3% increase in PRISM score (Figure 3A). We found a significant negative relationship between PRISM and GA such that PRISM scores decrease by 3% with increased GA (β =−0.03 (95% CI [−0.03-0.02], p <0.001) (Figure 3B).

**Figure 3:**
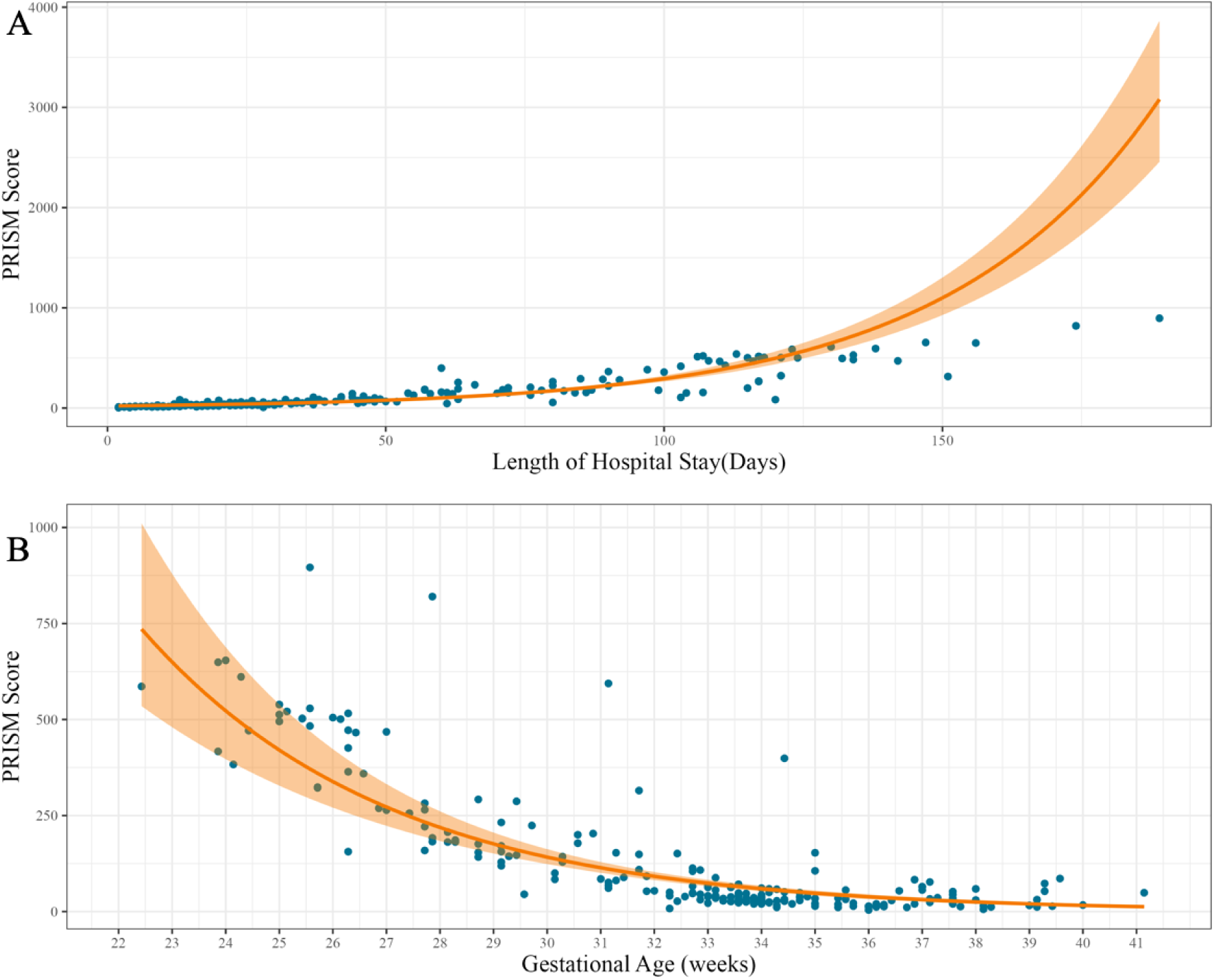
PRISM scores are significantly associated with GA and hospital length of stay. **A.** PRISM scores are positively associated with an infant’s length of stay in the hospital (β = 0.03 (95% CI [0.02, 0.03), p < 0.001). **B.** PRISM scores are negatively associated with an infant’s GA (β =−0.03 (95% CI [−0.03-0.02], p <0.001).

## 4. Discussion

Invasive respiratory interventions save infants’ lives. Unfortunately, these life-saving procedures have negative long term consequences for the preterm infant’s respiratory^8,39^ and neurological development^9,10,19,20^. It is crucial that we are able to accurately quantify and assess the respiratory interventions that infants receive throughout the entirety of their NICU admission to inform clinical decisions and provide realistic prognoses. A variety of tools have been developed to assess immediate respiratory needs, such as the likelihood of successful extubation^23^ or the appropriate fraction of inspired oxygen to deliver^22^. To our knowledge, however, no tool has been able to efficiently summarize the interventions needed to support an infant of any GA or respiratory need throughout their entire hospital stay. PRISM was designed with both clinicians and researchers in mind as a summative tool of respiratory health in the NICU. It accurately quantifies the invasive nature of respiratory support, and may serve as a useful tool to predict later neurodevelopmental dysfunction.

Most existing scores of infant respiratory outcomes use the most severe or chronic outcomes of BPD or CLD, conditions most prevalent in extremely premature infants^40^ who do not represent the majority of hospitalized infants in the NICU per year^41^. We tested PRISM’s ability to classify a diagnosis as RDS, CLD, or NoDx to determine its discriminative ability and potential future use as a diagnostic tool. Overall, PRISM was accurate at classifying infants who received respiratory diagnoses. It had perfect accuracy for identifying the most common respiratory disorder (RDS) and was moderately accurate at predicting the most severe respiratory disorder (CLD). However, it is important to note that PRISM did not classify any infant in the training set as receiving NoDx. For each respiratory disorder, precision values were above 0.90 but recall was quite low, indicating that PRISM is a conservative model where the classifications that it makes are correct, but it misses some positive cases. However, PRISM was trained on an imbalanced dataset with the majority of infants (n=155, 71%) receiving a diagnosis of RDS. Therefore, alternative statistics for ranking classifier performance, such as the AUC of each ROC curve can be particularly informative. PRISM’s micro-average AUC was above the clinical threshold of 0.75, indicating a potential for clinical value^42^. PRISM displays an ability to discriminate both across and within infants who receive respiratory diagnoses without the need for separate models for each diagnosis, a significant accomplishment given the subtle diagnostic differences between classes.

We also assessed PRISM’s performance against an existing, not-yet validated metric of neonatal respiratory function, the nSOFA-r score. Overall, the nSOFA-r score had 62% accuracy in distinguishing between diagnoses and was excellent at identifying RDS. However, the nSOFA-r’s ability to classify infants as having CLD was below chance. Much like PRISM, the nSOFA-r was unable to accurately identify cases in which the infant received no diagnosis.

Overall the Precision, Recall, and F1 score for both RDS and CLD were lower than those observed when using PRISM. AUC values for the NoDx class, RDS, and the macro-average fall well below the threshold for clinical utility. Only for the most severe respiratory diagnosis (CLD) and the micro-average does the nSOFA-r model reach AUC thresholds that are potentially clinically actionable. According to Delong’s Test PRISM outperforms nSOFA-r in its ability to stratify infants according to their likelihood of developing a specific respiratory diagnosis (CLD p < 0.001 and RDS p = 0.002).

We also assessed PRISM’s ability to identify diagnoses that commonly co-occur with invasive ventilation and respiratory dysfunction. PRISM identified cases of ROP with 95% accuracy, and IVH with 83% accuracy. Identification of ROP and IVH early in disease progression could facilitate the initiation of prophylactic treatment. PRISM’s unique ability to identify these disorders could support future work aiming to identify potential causative mechanisms related to distinct ventilation types and durations.

To further validate PRISM’s clinical utility, we assessed its association with metrics that have well known connections with overall respiratory health such as GA and length of hospital stay. Infants with a low GA and a long hospital stay are more likely to require intensive mechanical ventilation^21,31,32,43,44^, develop severe respiratory disorders^8,39^, and experience negative neurodevelopmental outcomes^9,10,19,20^. A gamma regression model showed a highly significant negative relationship between GA and PRISM, such that for each day that GA increases, PRISM decreases by 3%. Additionally, we found that PRISM was positively associated with the length of time that an infant spends in the NICU. While duration of ventilation is a factor included in the generation of PRISM scores, duration itself does not include the same variety of experience captured by PRISM. There are many other reasons that infants stay in the hospital for long periods of time aside from respiratory distress^45^. In fact, in this sample, infants spent an average of 23 days in the hospital on entirely room air and 67% of their stay consisted of no ventilation.While PRISM aligns with established predictors of an infant’s general health status it captures more fine grained health information, increasing its variation, and thus it’s accuracy.

### 4.1. Strengths and Limitations

The majority of existing studies which attempt to define respiratory or neurodevelopmental outcomes focus on infants born very or extremely premature^10^ or at low birth weights^9^, thus limiting generalizability, as 80% of NICU admissions have a GA above 34 weeks^41^ and in 2022 only 1.36% of live births were born at a very low birth weight^46^. One of the strengths of this study is that it includes infants of all GAs and is therefore more applicable to general NICU populations.

One limitation is that the sample was initially recruited to take part in a longitudinal neurodevelopmental study which required stable health status. As such, the sample does not represent some of the most severely ill infants who may have experienced the most invasive forms of ventilation. Finally, this study assesses PRISM’s clinical utility at only one site. Future studies should adopt a multisite approach to assess the generalizability of PRISM’s predictive performance across hospitals and neonatal populations.

## 5. Conclusion

Invasive ventilation measures have saved the lives of countless infants over the past 50 years; however, these machines are not without their consequences to an infant’s neurodevelopment and general health. This paper introduces PRISM as a tool to identify the long term consequences of ventilation. PRISM accurately summarizes the course of respiratory interventions experienced by an infant during their time in the NICU, discriminates between common respiratory disorders, and predicts which infants will develop common comorbid disorders. PRISM is designed for clinicians and researchers alike to assess respiratory support and provides an optimal metric for predicting an infant’s future health outcomes.

## Data Availability

All data produced in the present study are available upon reasonable request to the authors

