## Supplementary Materials for "The Prognostic Respiratory Intensity Scoring Metric (PRISM)"

### Supplement

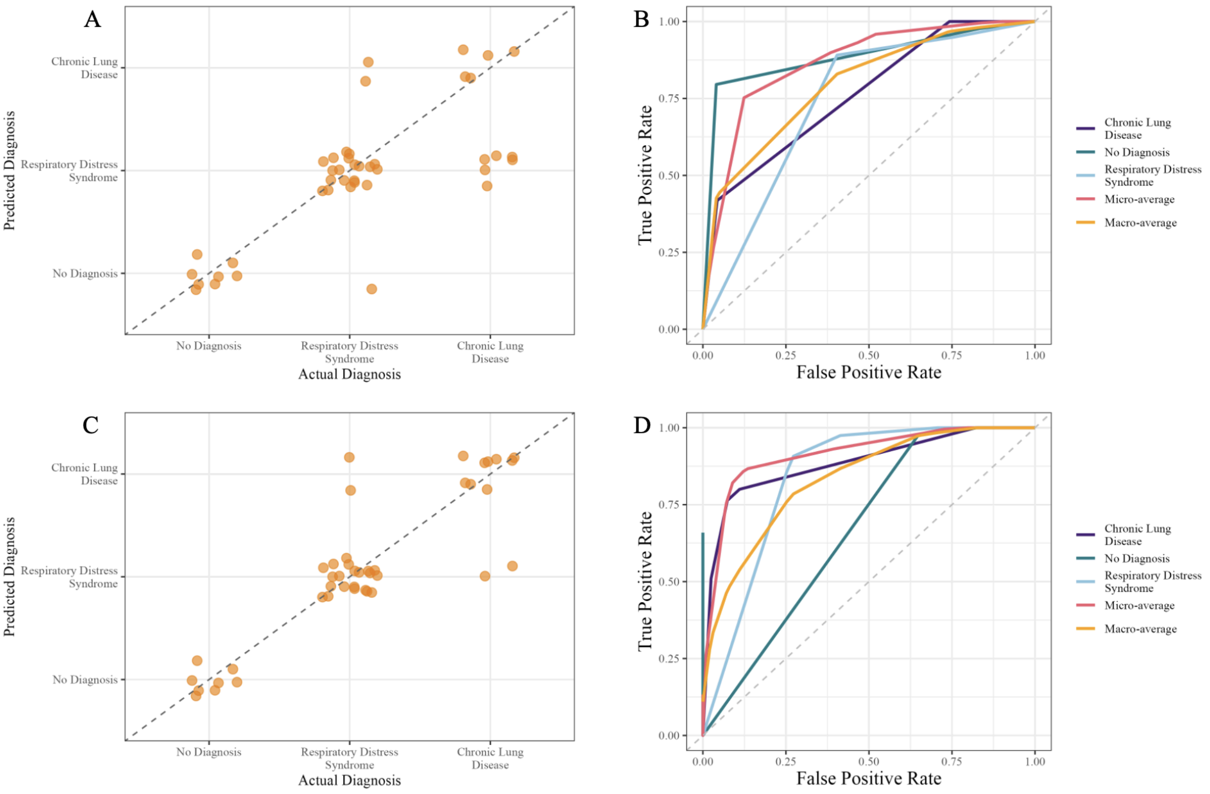


**Figure S1. PRISM-1 and PRISM-5 are highly accurate at predicting the incidence of the most common respiratory disorders in the NICU. A.** A CART model was trained on PRISM-1 scores to predict an infant's respiratory diagnosis. A scatter plot of predicted and actual diagnosis visualizes the 79% accuracy of the model. **B.** The performance of the CART model trained with PRISM-1 to classify RDS, CLD, and NoDx as well as the micro and macro-average curves were visualized with an ROC analysis. **C.** A CART model was trained on PRISM-5 scores to predict an infant's respiratory diagnosis. A scatter plot of predicted and actual diagnosis demonstrates the 91% accuracy for the model. **D.** The performance of the CART model trained with PRISM-1 to classify RDS, CLD, and NoDx as well as the micro and macro-average curves were visualized with an ROC analysis.

To determine the ability of PRISM scores to not only classify but predict the incidence of respiratory disorders in the NICU, we computed PRISM-1 and PRISM-5 which represent the PRISM scores on the first day of an infant’s hospital stay (PRISM-1) through the fifth day of an infant’s hospital stay (PRISM-5).

To assess the ability of PRISM-1 and PRISM-5 to predict the incidence of the most commonly occurring respiratory disorders in the NICU,we used the CART modelling technique[1] as detailed in the methods section. Predicted class probabilities were generated for training and test datasets and the optimal classification thresholds were determined by computing Youden's J statistic[2] from the ROC curves of each class. We performed predictions on the test set and if class probability exceeded the assigned threshold then the class with the highest predicted possibility was assigned.

We assessed the performance of both CART models by calculating the accuracy, precision, recall, and F1 score, for each possible class along with the micro and macro-average F1 score, precision, and recall. To visualize the diagnostic performance of the models we conducted an ROC analysis for each individual class (one vs. all binary outcomes) as well as the macro and micro-average using the pROC package[3] in R[4] (Figure S1). For each ROC curve, we calculated the area under the curve (AUC) values with a 95% Confidence Interval (CI) using a 10,000 iteration bootstrap resampling technique.

The CART model trained on PRISM-1 to predict respiratory diagnoses achieved an accuracy of 79% with high accuracy for NoDx and RDS. However, PRISM-1 struggled to identify CLD with the accuracy falling below chance levels. ROC-optimized thresholds, precision, recall, and F1 scores can be seen in Table S1. The AUC for each respiratory diagnosis class, with the exception of RDS, was above the 0.75 threshold of clinical utility. However RDS was right at this threshold with an AUC of 0.74. The micro and macro average AUC showed great discriminative ability for PRISM-1. The cart model trained on PRISM-5 had an even higher accuracy at 91% with each individual accuracy falling above 80% . ROC-optimized thresholds, precision, recall, and F1 scores can be seen in Table S1. PRISM-5 had excellent discriminative ability with each AUC falling well above the clinical threshold. Both of these early PRISM score calculations were adept at facilitating the prediction of later respiratory diagnoses. However PRISM-5 was substantially better at predicting which infants would receive the most severe diagnosis of CLD.

| | Threshold | Precision | Recall | F1 Score | Accuracy |
|---|---|---|---|---|---|
| | PRISM-1 | | | | |
| No Diagnosis | 0.44 | 1.00 | 0.97 | 0.99 | 100% |
| Respiratory Distress Syndrome | 0.45 | 0.81 | 0.68 | 0.74 | 87% |
| Chronic Lung Disease | 0.50 | 0.83 | 0.94 | 0.88 | 45% |
| Micro-Average | | 0.89 | 0.89 | 0.89 | |
| Macro-Average | | 0.88 | 0.86 | 0.89 | |
| | AUC | | | | |
| | PRISM-1 | | | | |
| No Diagnosis | 0.88 [0.82, 0.94] | | | | |
| Respiratory Distress Syndrome | 0.74, 0.68, 0.80] | | | | |
| Chronic Lung Disease | 0.76 [0.71, 0.82] | | | | |
| Micro-Average | 0.87 [0.83, 0.90] | | | | |
| Macro-Average | 0.80 [0.75, 0.84] | | | | |
| | Threshold | Precision | Recall | F1 Score | Accuracy |
| | PRISM Score -5 | | | | |
| No Diagnosis | 0.57 | 1.00 | 1.00 | 1 | 100% |
| Respiratory Distress Syndrome | 0.56 | 0.90 | 0.90 | 0.90 | 91% |
| Chronic Lung Disease | 0.19 | 0.93 | 0.90 | 0.92 | 82% |
| Micro-Average | | 0.95 | 0.94 | 0.95 | |
| Macro-Average | | 0.94 | 0.93 | 0.94 | |
| | AUC | | | | |
| | PRISM Score -5 | | | | |

| | |
|---|---|
| No Diagnosis | 0.89 [0.82, 0.93] |
| Respiratory Distress Syndrome | 0.84 [0.79, 0.89] |
| Chronic Lung Disease | 0.89 [0.83, 0.93] |
| Micro-Average | 0.91 [0.88, 0.94] |
| Macro-Average | 0.87 [0.83, 0.91] |

**Table S1: Model Fit Statistics for CART models trained with PRISM-1 and PRISM-5**
